# Geographic Heterogeneity in the Association Between Schooling and Cognitive Change Among Older Adults in India

**DOI:** 10.64898/2026.10.27.26366607

**Authors:** Priya Rajagopalan

## Abstract

Higher educational attainment is associated with steeper measured cognitive decline among older adults in India, but whether this association is uniform across the country is unknown. Using the Harmonized Diagnostic Assessment of Dementia for the Longitudinal Aging Study in India (wave 1, 2017–2019; wave 2, 2022–2024), this study examined whether the association between years of schooling and cognitive change varied across 18 states and union territories among 2,510 adults aged 60 years or older who were assessed at both waves. Models adjusted for age, sex, household wealth, and rural or urban residence. Change-score models related inter-wave change to schooling, and analysis of covariance additionally conditioned on baseline cognition. The schooling-by-geography interaction was significant in both specifications (change score, *F* = 2.97, *P* < .001; analysis of covariance, *F* = 4.11, *P* < .001). State-specific change-score estimates ranged from −0.056 SD per year of schooling in Odisha to +0.021 SD in Kerala. The pooled association reversed sign with baseline adjustment, from −0.022 SD (95% confidence interval: −0.027, −0.016) to +0.031 SD (95% confidence interval: 0.024, 0.038). A single national coefficient conceals substantial geographic variation in how schooling relates to cognitive aging in India.

## INTRODUCTION

Low educational attainment is among the most prominent modifiable risk factors for dementia worldwide.^1^ Its association with cognitive level in later life is strong and consistent across settings,^2,3^ but its association with the rate of cognitive change is much weaker and less consistent. A systematic review and meta-analysis of longitudinal cohorts found that education’s association with change was negligible and at least an order of magnitude smaller than its association with level,^4^ and an analysis of more than 170,000 adults in 33 Western countries found that education did not protect against age-related memory decline.^5,6^ These findings fit a passive account of cognitive reserve, in which education raises the starting point of the cognitive trajectory without changing its slope.^7,8^ Nearly all of this evidence, however, comes from high-income countries, and the few comparisons that span different national or institutional contexts show that the association between education and cognitive decline differs in strength and even in direction across settings.^9–11^

India sharpens this question. Using the Harmonized Diagnostic Assessment of Dementia for the Longitudinal Aging Study in India (LASI-DAD), Gross et al. reported an average decline in general cognitive performance of 0.029 SD per year, with steeper measured decline among more educated participants.^12^ Nichols et al. then examined that association directly with linear mixed models and found that participants with any schooling had higher baseline cognition but declined faster than those with none, a pattern that neither practice effects nor selective survival explained.^13^ That study examined variation by urbanicity, age, and gender, but not by state. The national association between schooling and decline in this cohort is therefore established; whether it holds uniformly across India is not.

There are good reasons to expect that it does not. Indian states differ in the timing and reach of mass schooling, in language, in household wealth, and in the burden of dementia, and LASI studies have documented marked interstate variation in both dementia prevalence and educational attainment.^14,15^ In the United States, the quality and resourcing of schooling in the state where people grew up are associated with later-life cognition beyond the number of years completed,^16–18^ and in Europe, inequality of educational opportunity at the time of schooling predicts cognitive functioning decades later.^19^ A year of schooling may therefore carry different cognitive meaning in different places. If it does, a single national coefficient describes no state well, and the inconsistent international findings on education and cognitive decline may partly reflect genuine contextual heterogeneity rather than methodological differences alone. No published study appears to have tested whether the association between schooling and cognitive change varies across the subnational units of a low- or middle-income country under a common protocol and a harmonized outcome. LASI-DAD, which applies one harmonized cognitive battery across 18 states and union territories, makes that test possible.^20,21^

A second, methodological issue shapes the analysis. Change-score and baseline-adjusted models estimate different quantities, and they can disagree in sign when exposure groups differ at baseline, a phenomenon known as Lord’s paradox.^22^ In nonrandomized data, neither model is unbiased in general. Analysis of covariance (ANCOVA) assumes that groups with the same baseline score would have changed alike, whereas the change score assumes that baseline differences would have persisted unchanged in the absence of exposure.^23^ Because schooling is completed decades before baseline and shapes baseline cognition, conditioning on baseline adjusts for a consequence of the exposure and can induce a spurious association, particularly when baseline is measured with error;^24^ change scores, for their part, do not estimate causal effects without strong assumptions.^25^ Regression to the mean is a related but separate concern: it arises from measurement error in the baseline score and can make change appear to depend on any characteristic correlated with baseline. This study had two aims: to test whether the association between schooling and cognitive change varies across Indian states and union territories, and to determine whether any such heterogeneity holds under both model specifications.

## METHODS

### Study population

LASI-DAD is an in-depth study of late-life cognition and dementia among adults aged 60 years or older, drawn from the nationally representative Longitudinal Aging Study in India by stratified sampling that oversampled participants at high risk of cognitive impairment.^20^ This analysis used wave 1 (2017–2019) and wave 2 (2022–2024); the wave 2 design is described elsewhere.^26^ Cognitive data came from Harmonized LASI-DAD release B.2 and household economic measures from Harmonized LASI release A.3, and records were linked on the participant identifier with a 100% match rate. Of 4,096 participants with wave 1 cognition and schooling, 2,511 had cognition at both waves and complete data on age, sex, state, and a recorded caste response, including “do not know.” Principal models, which additionally required household wealth and rural or urban residence, included 2,510 participants. Eighteen states and union territories, including the National Capital Territory (NCT) of Delhi (n = 74), met the inclusion threshold of at least 30 participants observed at both waves with nonzero variation in schooling. Uttarakhand appears under its former name, Uttaranchal, as labeled in the harmonized data.

### Cognitive outcome

The outcome was the harmonized general cognitive performance factor score at each wave, derived from the LASI-DAD battery by confirmatory factor analysis, with higher scores indicating better performance.^12,21^ The battery was administered in 10 languages,^20^ and the harmonized general factor has shown high marginal reliability in cross-national comparisons.^27^ Change was the wave 2 score minus the wave 1 score, so negative values indicate lower performance at follow-up. Mean follow-up was 4.7 years (SD, 0.7; range, 2.8 to 6.5 years), and unit means ranged from 3.9 to 5.7 years (Supplementary Table S3). Coefficients are expressed in SD units of the score per additional year of schooling over the observed interval, not per year of follow-up.

### Exposure and covariates

The exposure was completed years of schooling, modeled continuously. Principal models adjusted for age, sex, household wealth (untransformed household net assets), and rural or urban residence. Caste and marital status were treated as sensitivity covariates. In the 2,502 participants with a substantive caste category, which excluded 8 participants recorded as “do not know,” adding caste to the principal model changed the schooling coefficient from −0.0215 to −0.0217 SD, a shift of about 0.0002 SD. Because wealth and residence may partly reflect earlier education, the estimates are conditional associations rather than better-controlled total effects.^24^ Parental education was missing for all participants in the merged data and could not be included. Baseline state of residence defined the geographic unit.

### Statistical analysis

Two specifications described distinct associations. The change-score model related inter-wave change to schooling and covariates, whereas ANCOVA related wave 2 cognition to schooling, baseline cognition, and the same covariates. The change score served as the principal description because it summarizes observed within-person change without conditioning on a baseline score that schooling helped to produce;^24^ it was not assumed to estimate a causal effect.^25^ Comparing the two schooling coefficients shows sensitivity to baseline conditioning; it neither diagnoses regression to the mean nor selects a causal estimator.^22,23^ With only 2 waves, a linear mixed model with a schooling-by-time term draws on the same within-person information as the change score, so the longitudinal model described below served as a matched check rather than as independent evidence.

Geographic heterogeneity was the primary question. It was tested with a joint schooling-by-geography interaction, comparing a model with geography indicators and their interactions with schooling against a model with a common schooling slope, in both the change-score and ANCOVA specifications. This fixed-effects test treats the 18 geographic units as those observed rather than as a sample from a larger population. It asks whether schooling slopes differ across units; it does not establish their direction, identify which units differ, or validate a ranking. A matched longitudinal likelihood-ratio test of the three-way schooling-by-time-by-geography interaction compared linear mixed models with and without that term on identical rows (2,510 participants; 5,020 person-waves) under the principal adjustment set, estimated by maximum likelihood. Two secondary, descriptive summaries were also used. The first was a random-slope mixed model; because the slope variance lies on the boundary of its parameter space under the null hypothesis, its likelihood-ratio statistic was referred to a 50:50 mixture of χ^2^ distributions with 1 and 2 degrees of freedom.^28^ The second was a random-effects meta-analysis of unit-specific slopes from separate within-unit regressions, with the between-unit variance estimated by restricted maximum likelihood, the pooled confidence interval and prediction interval derived by the Knapp-Hartung method (*t* distribution, *k* − 1 degrees of freedom), and heterogeneity summarized by the *I*^2^ statistic.^29–32^ The prediction interval was reported because it describes the range of associations expected in a comparable unit, which *I*^2^ does not.^33,34^ These summaries share participants with the principal models and are not independent replications.

Robustness was assessed in several ways. Stabilized inverse-probability-of-censoring weights, truncated at the 1st and 99th percentiles, modeled return at wave 2 as a function of baseline cognition, schooling, age, sex, household wealth, residence, and geographic unit.^35^ Because LASI-DAD oversampled participants at high risk of cognitive impairment,^20^ a design-weighted check used the baseline respondent weight; full survey inference with strata and primary sampling units was not completed and is noted as a limitation.^36^ A model adding diabetes and body mass index, both from the LASI baseline interview, was compared with the principal model fitted to the same 2,408 participants with both measures. The interaction tests were repeated with adjustment for the length of follow-up and its interaction with schooling, with annualized change as the outcome, with HC3 standard errors, after excluding the 3 units with fewer than 100 participants, and with age, sex, and wealth coefficients allowed to differ by unit. A leave-one-unit-out procedure re-estimated the pooled slope and the interaction test after dropping each unit in turn. All other ordinary least squares models used HC1 heteroskedasticity-consistent standard errors. Analyses were conducted in R, version 4.4.1 (R Foundation for Statistical Computing, Vienna, Austria), with the lme4 package, version 1.1-35.5, for mixed models^37^ and the metafor package, version 4.6-0, applied to the aggregate unit estimates for the meta-analytic summaries.^38^

Analysis code is deposited on Zenodo (https://doi.org/10.5281/zenodo.23172557) and will be released on publication; an anonymized copy is provided for review.

This secondary analysis used de-identified data obtained under the LASI-DAD data use agreement and analyzed within a secure data enclave. The parent study received ethics approval, and all participants gave informed consent, as described in the LASI-DAD design papers.^20,26^ Because the present study used only de-identified secondary data, it did not constitute human-subjects research and did not require additional institutional review board approval.

## RESULTS

### Sample characteristics

The analytic sample included 2,511 adults across 18 states and union territories (mean age, 68.2 years [SD, 6.3]; 56.1% women). Mean schooling was 3.9 years, and almost half of participants reported no formal schooling. Baseline cognition rose steeply with education, from −0.50 SD in the no-schooling group to 0.96 SD among those with at least 6 years (Table 1). Unadjusted change followed the opposite gradient, from −0.01 SD among those with no schooling to −0.27 SD among those with at least 6 years; these values describe change over the observed interval and are distinct from differences in baseline level. Unit samples ranged from 71 participants in Madhya Pradesh to 212 in Kerala, and attrition, schooling, and baseline cognition varied across units (Supplementary Table S3).

**Table 1.** Baseline characteristics of the analytic sample by educational attainment, LASI-DAD waves 1-2 (2017-2024)

| Characteristic | Overall | No schooling | Primary (1-5) | Secondary+ |
| --- | --- | --- | --- | --- |
| No. | 2,511 | 1,207 | 505 | 799 |
| Age, years, mean (SD) | 68.2 (6.3) | 68.5 (6.7) | 68.6 (6.2) | 67.3 (5.7) |
| Female, % | 56.1 | 73.7 | 49.5 | 33.7 |
| Baseline cognition, SD | 0.10 (0.90) | −0.50 (0.60) | 0.15 (0.66) | 0.96 (0.66) |
| Change in cognition, SD | −0.12 (0.62) | −0.01 (0.63) | −0.15 (0.59) | −0.27 (0.58) |
| Follow-up, years | 4.65 (0.66) | 4.63 (0.62) | 4.64 (0.68) | 4.70 (0.70) |
| Schooling, years | 3.88 (4.63) | 0.00 (0.00) | 3.69 (1.22) | 9.87 (2.76) |
| Scheduled caste, % | 18.6 | 25.3 | 13.3 | 11.8 |
| Scheduled tribe, % | 5.2 | 6.4 | 5.1 | 3.4 |
| Other backward class, % | 41.8 | 41.8 | 44.0 | 40.4 |
| No caste or other, % | 34.1 | 26.1 | 37.2 | 44.3 |
| Urban residence, % | 34.4 | 22.5 | 36.0 | 51.3 |
| Rural residence, % | 65.6 | 77.5 | 64.0 | 48.7 |

Abbreviation: SD, standard deviation. Values are mean (SD) unless marked %. Education groups: none (0 years), primary (1–5 years), and secondary or higher (6 or more years). Caste included a labeled “do not know” category (0.3% overall). Group differences were significant for all rows except follow-up time ( $P$ from Kruskal-Wallis or $\chi^2$ tests).

### The sign of the pooled association depends on the model

In the principal change-score model, each additional year of schooling was associated with −0.022 SD of change (95% CI: −0.027, −0.016; N = 2,510), so a 10-year difference in schooling corresponded to a 0.22 SD larger decline over the interval, nearly twice the mean change of −0.12 SD. In ANCOVA on the same participants, the coefficient was +0.031 SD (95% CI: 0.024, 0.038). A covariate ladder located the reversal in the baseline term: distal covariates barely moved the change-score estimate, whereas adding baseline cognition reversed its sign (Table 2).

**Table 2.** Schooling and cognitive change under two models, with a covariate ladder.

| Model | Estimate | HC1 SE | 95% CI |
| --- | --- | --- | --- |
| Change score (principal) | −0.022 | 0.003 | −0.027, −0.016 |
| Change score + state fixed effects | −0.015 | 0.003 | −0.020, −0.009 |
| ANCOVA (principal) | 0.031 | 0.004 | 0.024, 0.038 |
| ANCOVA + state fixed effects | 0.026 | 0.003 | 0.019, 0.032 |
| Covariate ladder (change score): |  |  |  |
| Age + sex | −0.028 | 0.003 | −0.034, −0.023 |
| Age + sex + caste | −0.028 | 0.003 | −0.033, −0.023 |
| Principal + baseline cognition | 0.031 | 0.004 | 0.024, 0.038 |
| Age + sex + residence | −0.021 | 0.003 | −0.027, −0.016 |
| Principal + diabetes and BMI (n = 2,408) | −0.021 | 0.003 | −0.027, −0.015 |

Abbreviations: ANCOVA, analysis of covariance; BMI, body mass index; CI, confidence interval; HC1, heteroskedasticity-consistent standard error, type 1; SD, standard deviation; SE, standard error. Estimates are change in cognition per additional year of schooling, in SD units. Principal models (N = 2,510) adjust for age, sex, household wealth, and rural or urban residence; caste and marital status are sensitivity covariates. The covariate-ladder rows are separate modifications of a base model, not a single cumulative sequence; the diabetes and BMI row adds both measures to the principal model in the 2,408 participants with both available. Adding baseline cognition to the change-score model reverses the sign; this is the baseline-conditioning contrast, not proof of regression to the mean.

Because this contrast reflects baseline conditioning without identifying its cause,^23,24^ the remaining analyses report both models rather than choosing between them.

### The association varies across states and union territories

The association between schooling and cognitive change was not constant across Indian states and union territories. The schooling-by-geography interaction was significant in both specifications, with HC1-robust joint tests giving *F* = 2.97 for the change-score model (df = 17, 2,470; *P* = 4 × 10^−5^) and *F* = 4.11 for ANCOVA (df = 17, 2,469; *P* = 3 × 10^−8^) (Table 3). The matched longitudinal test of the three-way schooling-by-time-by-geography interaction led to the same conclusion (likelihood-ratio χ^2^ = 57.5; df = 17; *P* = 3 × 10^−6^), indicating that the change in the schooling gradient over time differed across geographic units. A random schooling slope also improved fit over a random-intercept model (likelihood-ratio χ^2^ = 32.1; boundary-corrected *P* = 6 × 10^−8^), with an estimated between-unit SD of the schooling slope of 0.014.

**Table 3.**
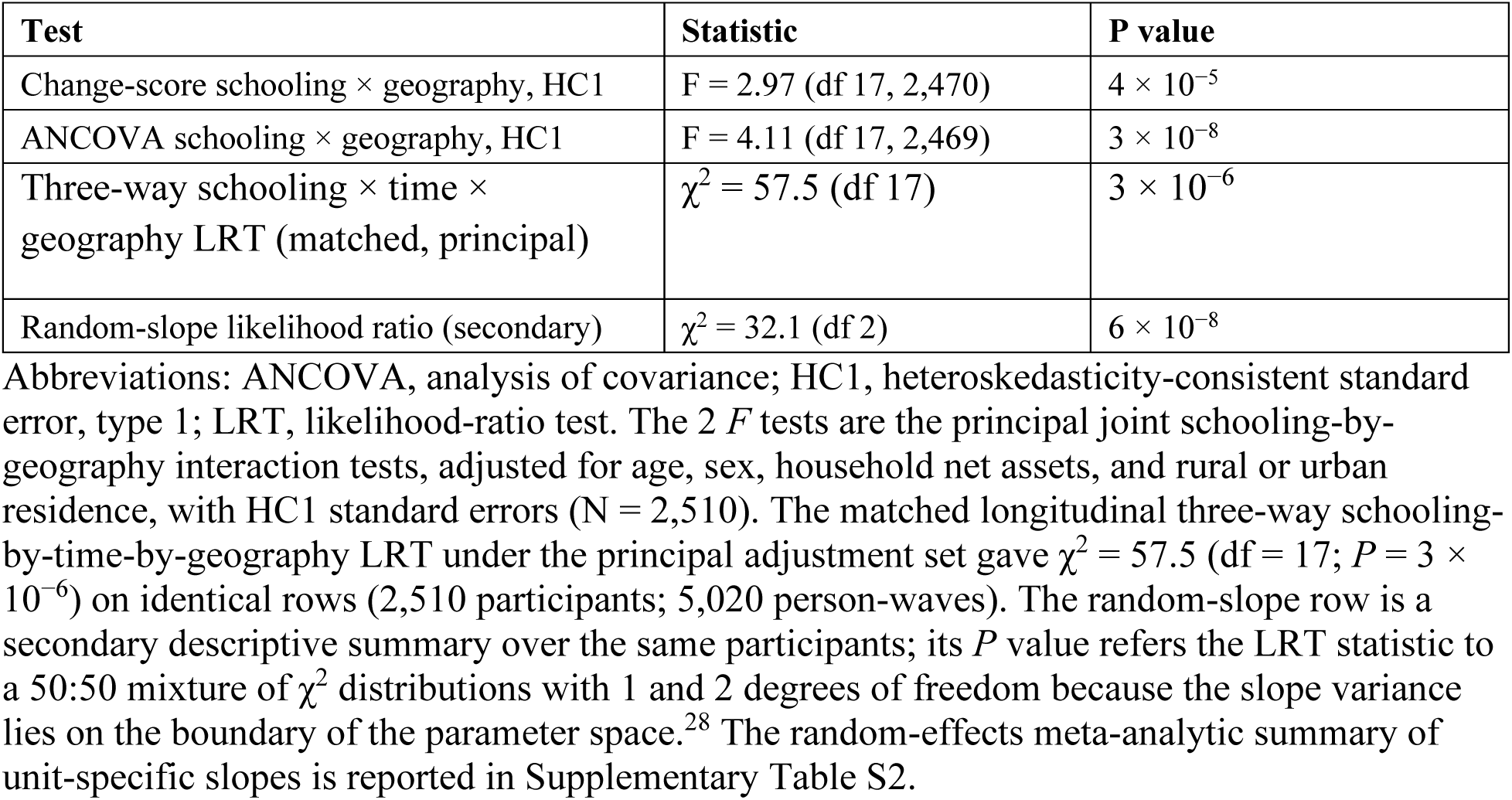
Tests of geographic heterogeneity in the schooling-cognition association (principal adjustment set)

| Test | Statistic | P value |
| --- | --- | --- |
| Change-score schooling × geography, HC1 | $F = 2.97$ (df 17, 2,470) | $4 \times 10^{-5}$ |
| ANCOVA schooling × geography, HC1 | $F = 4.11$ (df 17, 2,469) | $3 \times 10^{-8}$ |
| Three-way schooling × time × geography LRT (matched, principal) | $\chi^2 = 57.5$ (df 17) | $3 \times 10^{-6}$ |
| Random-slope likelihood ratio (secondary) | $\chi^2 = 32.1$ (df 2) | $6 \times 10^{-8}$ |

Abbreviations: ANCOVA, analysis of covariance; HC1, heteroskedasticity-consistent standard error, type 1; LRT, likelihood-ratio test. The 2 $F$ tests are the principal joint schooling-by-geography interaction tests, adjusted for age, sex, household net assets, and rural or urban residence, with HC1 standard errors ( $N = 2,510$ ). The matched longitudinal three-way schooling-by-time-by-geography LRT under the principal adjustment set gave $\chi^2 = 57.5$ (df = 17; $P = 3 \times 10^{-6}$ ) on identical rows (2,510 participants; 5,020 person-waves). The random-slope row is a secondary descriptive summary over the same participants; its $P$ value refers the LRT statistic to a 50:50 mixture of $\chi^2$ distributions with 1 and 2 degrees of freedom because the slope variance lies on the boundary of the parameter space.<sup>28</sup> The random-effects meta-analytic summary of unit-specific slopes is reported in Supplementary Table S2.

In the principal pooled interaction model, unit-specific estimates ranged from −0.056 SD per additional year of schooling in Odisha (95% CI: −0.078, −0.034) to +0.021 SD in Kerala (95% CI: 0.006, 0.036). In Odisha, a 10-year difference in schooling thus corresponded to a 0.56 SD larger decline, more than 4 times the mean change in the cohort, whereas in Kerala it corresponded to a 0.21 SD smaller decline. These state-specific estimates are mapped in Supplementary Figure S3. The association was negative in 17 of the 18 units, and confidence intervals excluded zero in 8 units: 7 negative (NCT Delhi, Gujarat, Haryana, Maharashtra, Odisha, Telangana, and Uttaranchal) and 1 positive, Kerala (Table 4, Figure 1). In the ANCOVA specification, 12 units had positive intervals that excluded zero and none had an entirely negative interval (Supplementary Table S1, Supplementary Figure S1). These pointwise intervals are not corrected for multiple comparisons and are reported for completeness rather than as a ranking.

**Figure 1.**
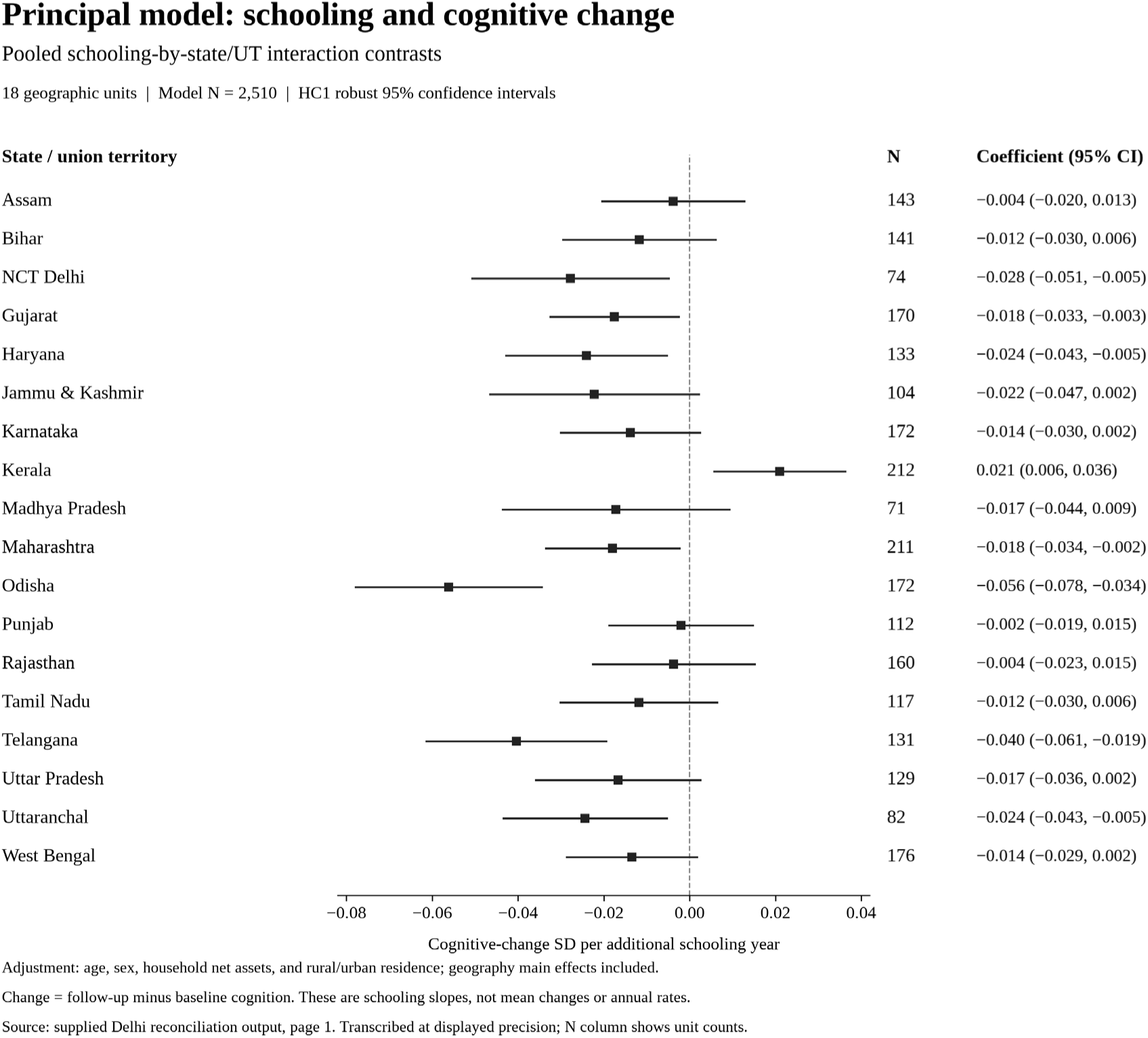
State- and union-territory-specific associations between schooling and cognitive change from the principal pooled interaction model, Longitudinal Aging Study in India–Diagnostic Assessment of Dementia, 2017–2024. Estimates are adjusted for age, sex, household net assets, and rural or urban residence, with geography main effects. Squares are unit-specific contrasts, and horizontal lines are 95% confidence intervals (coefficient ± 1.96 × HC1 standard error). The dashed line marks zero. The association is negative in 17 of 18 units and positive in Kerala. The principal evidence for heterogeneity is the joint interaction test in Table 3, *F*(17, 2,470) = 2.97, *P* = 4 × 10^−5^; pointwise intervals are not corrected for multiple comparisons. HC1, heteroskedasticity-consistent standard error, type 1.

**Table 4.**
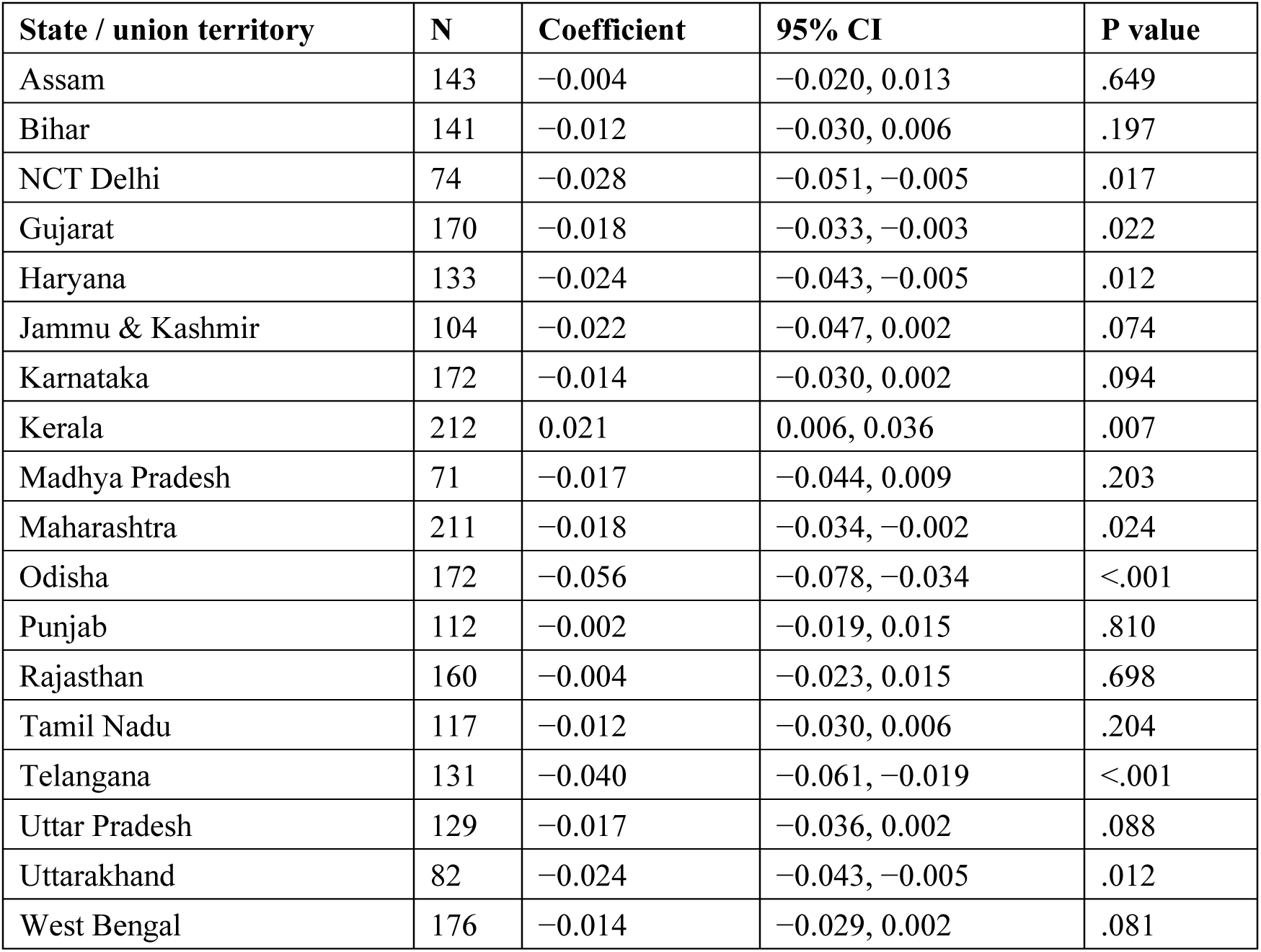
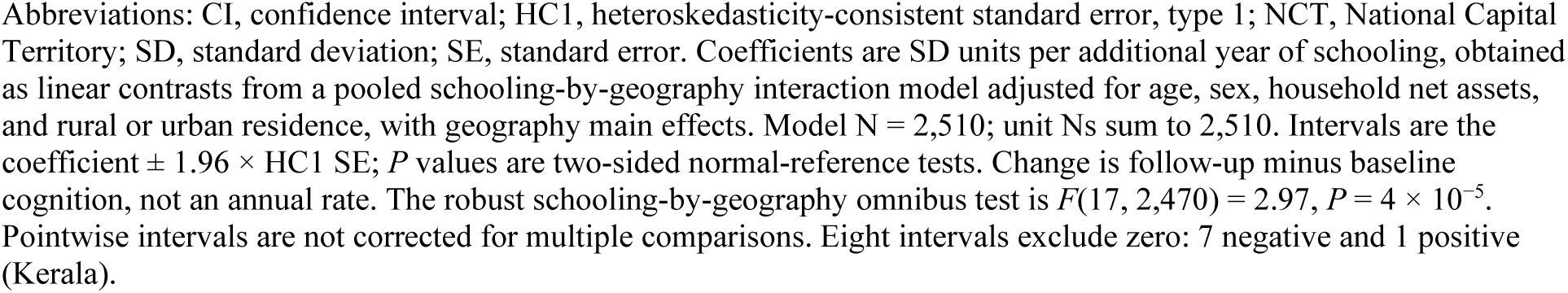
State- and union-territory-specific schooling associations with cognitive change (principal pooled interaction model)

| State / union territory | N | Coefficient | 95% CI | P value |
| --- | --- | --- | --- | --- |
| Assam | 143 | -0.004 | -0.020, 0.013 | .649 |
| Bihar | 141 | -0.012 | -0.030, 0.006 | .197 |
| NCT Delhi | 74 | -0.028 | -0.051, -0.005 | .017 |
| Gujarat | 170 | -0.018 | -0.033, -0.003 | .022 |
| Haryana | 133 | -0.024 | -0.043, -0.005 | .012 |
| Jammu & Kashmir | 104 | -0.022 | -0.047, 0.002 | .074 |
| Karnataka | 172 | -0.014 | -0.030, 0.002 | .094 |
| Kerala | 212 | 0.021 | 0.006, 0.036 | .007 |
| Madhya Pradesh | 71 | -0.017 | -0.044, 0.009 | .203 |
| Maharashtra | 211 | -0.018 | -0.034, -0.002 | .024 |
| Odisha | 172 | -0.056 | -0.078, -0.034 | <.001 |
| Punjab | 112 | -0.002 | -0.019, 0.015 | .810 |
| Rajasthan | 160 | -0.004 | -0.023, 0.015 | .698 |
| Tamil Nadu | 117 | -0.012 | -0.030, 0.006 | .204 |
| Telangana | 131 | -0.040 | -0.061, -0.019 | <.001 |
| Uttar Pradesh | 129 | -0.017 | -0.036, 0.002 | .088 |
| Uttarakhand | 82 | -0.024 | -0.043, -0.005 | .012 |
| West Bengal | 176 | -0.014 | -0.029, 0.002 | .081 |

Abbreviations: CI, confidence interval; HC1, heteroskedasticity-consistent standard error, type 1; NCT, National Capital Territory; SD, standard deviation; SE, standard error. Coefficients are SD units per additional year of schooling, obtained as linear contrasts from a pooled schooling-by-geography interaction model adjusted for age, sex, household net assets, and rural or urban residence, with geography main effects. Model N = 2,510; unit Ns sum to 2,510. Intervals are the coefficient $\pm 1.96 \times \text{HC1 SE}$ ; $P$ values are two-sided normal-reference tests. Change is follow-up minus baseline cognition, not an annual rate. The robust schooling-by-geography omnibus test is $F(17, 2,470) = 2.97$ , $P = 4 \times 10^{-5}$ . Pointwise intervals are not corrected for multiple comparisons. Eight intervals exclude zero: 7 negative and 1 positive (Kerala).

Separate within-unit regressions on the same adjustment set, which allow covariate coefficients to differ by unit, gave a broadly similar descriptive picture (Supplementary Table S2, Supplementary Figure S2). The random-effects summary of these slopes was −0.014 SD (95% CI: −0.022, −0.007; *P* < .001), with moderate heterogeneity (*I*^2^ = 46%; *Q* = 31.5, df = 17; *P* = .02). The 95% prediction interval, −0.038 to +0.009, included zero, implying that in a comparable unit the association could plausibly range from clearly negative to null or weakly positive. In these separate regressions, 5 units had negative intervals that excluded zero (Haryana, Uttar Pradesh, Odisha, Karnataka, and Telangana) and none had a positive interval that did so; Kerala, Punjab, and Rajasthan had positive point estimates with intervals that included zero.

### Robustness

Participants observed at follow-up were younger (68.2 vs. 72.2 years) and had higher baseline cognition (0.10 vs. −0.20 SD) than those who were not. In the return model, baseline cognition predicted participation (logit coefficient, 0.58; *P* < .001), and schooling was inversely associated with it (−0.047; *P* < .001). With censoring weights, the schooling coefficient was −0.0213, compared with −0.0216 unweighted in the same participants (Table 5). The closeness of these estimates is consistent with, but does not prove, limited attrition bias with respect to the measured predictors. Selection remains a concern because LASI-DAD mortality analyses link dementia and cardiometabolic phenotypes to death.^39^

**Table 5.** Sensitivity analyses for the pooled schooling-cognition association.

| Analysis | Slope (SE) | No. |
| --- | --- | --- |
| Primary, unweighted | −0.0216 (0.0029) | 2,510 |
| Inverse-probability-of-censoring weighted | −0.0210 (0.0029) | 2,510 |
| Design-weighted, baseline respondent weight | −0.0218 (0.0030) | 2,510 |
| Principal + diabetes and BMI | −0.0209 (0.0029) | 2,408 |
| Leave-one-unit-out, schooling-by-geography F test | F range, 1.80 to 3.18 | Varies |
| Leave-one-unit-out, pooled slope | Range −0.023 to −0.019 | Varies |

Abbreviations: BMI, body mass index; SE, standard error. All rows use the principal adjustment set (age, sex, household net assets, and rural or urban residence). The stabilized censoring weight, truncated at the 1st and 99th percentiles, modeled return at wave 2 from baseline cognition, schooling, age, sex, household net assets, residence, and geographic unit. Unweighted and weighted estimates are close but not identical, which is consistent with, but not proof of, limited attrition bias. The design-weighted row uses the baseline respondent weight only; full survey inference with strata and primary sampling units is not included. The diabetes and BMI row is estimated in the 2,408 participants with both measures; the principal model in the same participants gives −0.0216. Leave-one-unit-out rows drop 1 unit at a time from the 2,510 participants.

Restricting the principal model to the 2,408 participants with diabetes and body mass index data left the schooling coefficient essentially unchanged (−0.0216 SD), and adding both measures moved it only slightly toward zero (−0.0209 SD; 95% CI: −0.0266, −0.0151). The 102 participants without both measures had slightly lower baseline cognition (−0.12 vs. 0.10 SD; P = .01) but did not differ in schooling (P = .92) and were similar in age (P = .10). Measured cardiometabolic burden therefore accounted for little of the association, although the link between diabetes and cognition in India differs from patterns in high-income countries and is itself modified by education.^40^ A design-weighted check using the baseline respondent weight returned a similar point estimate (−0.022 SD). Leave-one-unit-out analysis moved the pooled slope by at most 0.003 SD, with the largest shift on excluding Kerala, and the interaction remained significant whichever unit was dropped (F range, 1.80 to 3.18; largest P = .03; Table 5). The interaction tests also remained significant in every sensitivity analysis (all *P* < .001; Supplementary Table S4).

## DISCUSSION

This study shows that the association between schooling and cognitive change among older adults in India is not geographically uniform. The schooling-by-geography interaction was significant under both the change-score and the baseline-adjusted specification and in a matched longitudinal mixed model, and state-specific estimates spanned a wide range, from strongly negative in Odisha to positive in Kerala. Earlier LASI-DAD analyses established that more educated participants declined faster on average;^12,13^ the present results show that this national average combines states in which the association is strongly negative with at least one in which it is reversed. Characterizing that geographic structure, which had not previously been described in this cohort or, as far as published evidence shows, within any low- or middle-income country, is the principal contribution of this study.

These findings may help explain why the international evidence on education and cognitive decline is inconsistent. In high-income cohorts, education is related to cognitive level but only weakly, if at all, to the rate of decline.^2,4–6^ By contrast, education was associated with slower decline in China than in England,^10^ with delayed onset of accelerated decline in the Health and Retirement Study and its sister studies,^41^ and with slower or faster decline depending on the cognitive domain in a 14-cohort international consortium that included no cohort from India.^9^ In Europe, educational differences in cognitive decline vary by welfare regime.^11^ Because each of these comparisons crosses study designs, languages, and instruments, contextual heterogeneity cannot be separated from methodological difference. The present analysis holds the protocol and the harmonized outcome constant and still finds heterogeneity across states, which suggests that part of the international inconsistency reflects real differences in what schooling means for cognitive aging in different settings. The steeper decline among more educated Indians reported nationally^13^ is consistent with the negative association observed in most states. Under passive reserve accounts, education raises the level from which decline begins without altering its rate;^7^ under compensation accounts, the advantage it confers is depleted more rapidly once pathology accumulates, producing steeper measured decline among the more educated.^8,42^ Most states fit the second pattern, but in Kerala more schooling tracked slower decline, which suggests that the reserve mechanism may differ across settings and that longer or better schooling may build reserve that slows decline rather than only raising its starting point; a single national coefficient obscures that contrast.

Several features of Indian states could produce this pattern. Today’s older adults were schooled when the reach, quality, and language of instruction differed sharply across states, and early LASI data showed large interstate differences in educational attainment.^15^ Indicators of childhood school quality are associated with later-life cognition independently of years completed,^43^ and evidence from the United States indicates that the resources of state school systems shape later cognition beyond attainment,^17,18^ so a year of schooling may represent more durable cognitive capital in some states than in others. Interstate differences in survival and attrition, in cardiometabolic and environmental exposures, and in the burden of dementia^14^ could also contribute.

Kerala and Odisha, the 2 extremes, also differed in their schooling histories: mean schooling was 7.4 years in Kerala and 3.7 years in Odisha, 8% of the Kerala sample had no schooling compared with 38% in Odisha, and baseline cognition was higher in Kerala (Supplementary Table S3). Longer, earlier, and more uniform schooling may allow an added year to become cognitive capital that persists, but the 2 states also differed in attrition, and whether the contrast reflects school quality, selective survival, or measurement cannot be settled with these data. The study was designed to detect heterogeneity rather than to explain it, and these explanations should be tested directly with state-level measures of historical school quality and language of instruction.

The pooled sign was also model-dependent: negative in change scores and positive after baseline adjustment. That reversal should not be read as evidence that education protects cognition.

Glymour et al., using education and cognitive change as their example, showed that baseline adjustment can create a spurious association,^24^ and in nonrandomized comparisons the choice between ANCOVA and change scores rests on untestable assumptions about what would have happened without the exposure.^22,23^ Neither estimate is a causal effect.^25^ The safer reading is that studies of cognitive aging should report both the level and the change association and state how their conclusions depend on baseline conditioning. The geographic heterogeneity, however, did not depend on that choice, because the interaction was significant under both specifications.

Geography in this analysis is a marker, not a mechanism. Baseline state is an imperfect proxy for where schooling occurred, and because the LASI-DAD battery was administered in 10 languages,^20^ language of assessment is closely tied to state. Cognitive items in LASI show differential item functioning by education and literacy,^44^ and items transported across cultural contexts perform differently.^45^ If the measurement properties of the battery differ jointly by language and education, part of the observed heterogeneity could reflect measurement rather than cognitive aging. The harmonized factor score^21,27^ and the within-person change outcome reduce, but do not remove, that concern. Cross-sectional neuroimaging in this cohort shows that education, literacy, and rural residence modify brain–cognition associations,^46^ which is consistent with a contextual interpretation but does not establish a mechanism for state-specific change.

### Strengths and limitations

This study has several strengths: repeated cognitive assessment with a harmonized outcome in a large, geographically diverse national cohort; comparison of both model specifications in the same participants; and a heterogeneity test that does not rest on a superpopulation assumption, corroborated by a matched longitudinal test. Its limitations bound the claims. Two waves support inter-wave change but not trajectories, and they cannot fully separate aging, practice effects, and measurement error. Follow-up varied, so change is not an annual rate, although adjusting for the interval did not alter the conclusions. Schooling was self-reported and did not capture quality, and parental education was unavailable. With 18 units and as few as 71 participants in one of them, unit-specific intervals are wide: at 80% power and a 2-sided α of .05, a unit-specific slope would have to differ from zero by about 2.8 standard errors, roughly 0.021 SD in Gujarat and 0.038 SD in Madhya Pradesh, to be detected. Caste, which shapes both access to schooling and its returns, was treated as an adjustment variable rather than examined as a modifier. The complete-case sample need not retain the parent cohort’s representativeness, censoring weights cannot address unmeasured selection, and full survey-weighted inference was not completed. Air pollution and other environmental exposures were not modeled.^1^ Finally, these observational results do not establish that schooling causes faster or slower cognitive change.

These findings have two implications. For research, pooled estimates of cognitive aging in large and diverse countries should routinely be accompanied by a formal heterogeneity test and a prediction interval, because a precise national coefficient can coexist with materially different subnational associations.^33,34^ For policy, dementia prevention and cognitive health planning in India, where the burden of dementia is already known to be unevenly distributed across states,^14^ should not assume that the cognitive returns to schooling are the same everywhere.

## Conclusion

The association between schooling and cognitive change among older adults in India varies across states and union territories, and that variation holds under both change-score and baseline-adjusted models. The pooled association was robust to attrition weighting, design weighting, cardiometabolic adjustment, and the exclusion of any single unit, but its sign depended on baseline conditioning. National summaries of education and cognitive aging in India should therefore be interpreted alongside, not instead of, their geographic variation.

## Data Availability

Data analysis was conducted entirely within the secure Gateway to Global Aging Data Enclave, managed by the University of Southern California, where all operations were restricted to the secure environment and only aggregated, vetted statistical outputs were cleared for export.
This research uses harmonized variables provided by the Gateway to Global Aging Data platform (funded by National Institute on Aging grant R01 AG030153) and the core Longitudinal Aging Study in India Diagnostic Assessment of Dementia (LASI-DAD).
Access requires registration and a data use agreement through the Gateway to Global Aging Data (https://g2aging.org). Analysis code is deposited on Zenodo (doi:10.5281/zenodo.23172557) and will be released on publication.

https://doi.org/10.5281/zenodo.23172557

## Abbreviations

ANCOVA: analysis of covariance
CI: confidence interval
HC1: heteroskedasticity-consistent standard error: type 1
HC3: heteroskedasticity-consistent standard error: type 3
LASI: Longitudinal Aging Study in India
LASI-DAD: Harmonized Diagnostic Assessment of Dementia for the Longitudinal Aging Study in India
NCT: National Capital Territory
REML: restricted maximum likelihood
SD: standard deviation.

## Acknowledgments

Data were provided by the Harmonized Diagnostic Assessment of Dementia for the Longitudinal Aging Study in India (Harmonized LASI-DAD release B.2) and Harmonized LASI release A.3, accessed through the Gateway to Global Aging Data. The author used an artificial intelligence assistant (Perplexity Computer; Perplexity AI) for analysis-code drafting, numerical verification, and language editing; the author reviewed and verified all analyses and text and takes full responsibility for the content.

## Author contributions (CRediT)

P.R.: conceptualization, data curation, formal analysis, methodology, software, visualization, writing (original draft), and writing (review and editing). P.R. is the sole author.

## Data availability

LASI-DAD data are available to qualified researchers through the Gateway to Global Aging Data (https://g2aging.org) and are analyzed within a secure enclave; the author is not permitted to redistribute the data. Analysis code is deposited on Zenodo (https://doi.org/10.5281/zenodo.23172557) and cited in the reference list; it will be made public on publication, and an anonymized copy is provided as a supplementary file for review.

## Disclaimer

none.

## SUPPLEMENTARY MATERIAL

**Supplementary Figure S1.**
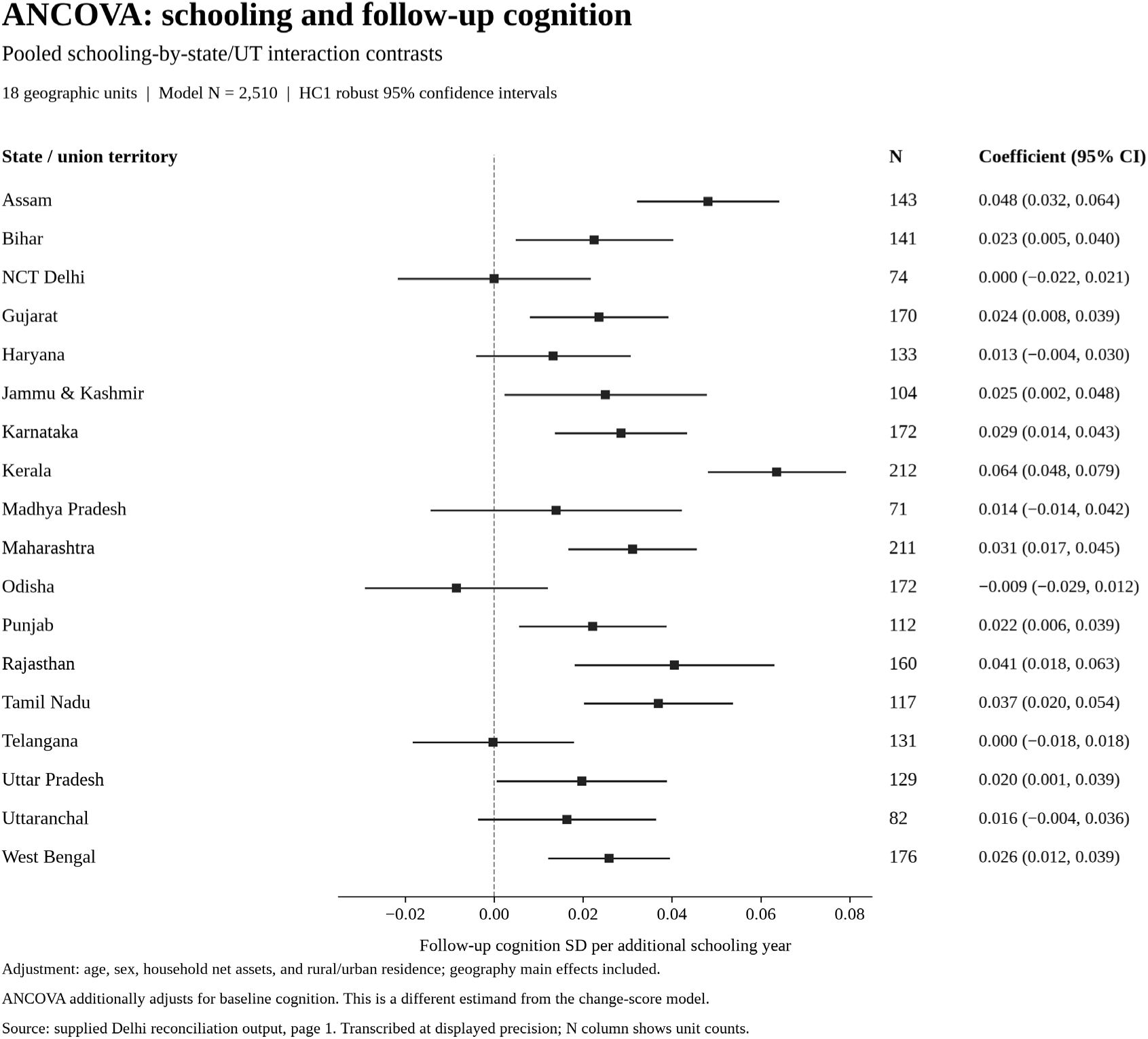
Forest plot of the ANCOVA pooled interaction contrasts (schooling association with follow-up cognition, conditional on baseline cognition), same adjustment set as Figure 1. *Forest plot of the ANCOVA pooled interaction contrasts (schooling association with follow-up cognition, conditional on baseline cognition), same adjustment set as* Figure 1*. This is a different estimand from the change-score model and is shown as a companion, not as confirmation*.

**Supplementary Figure S2.**
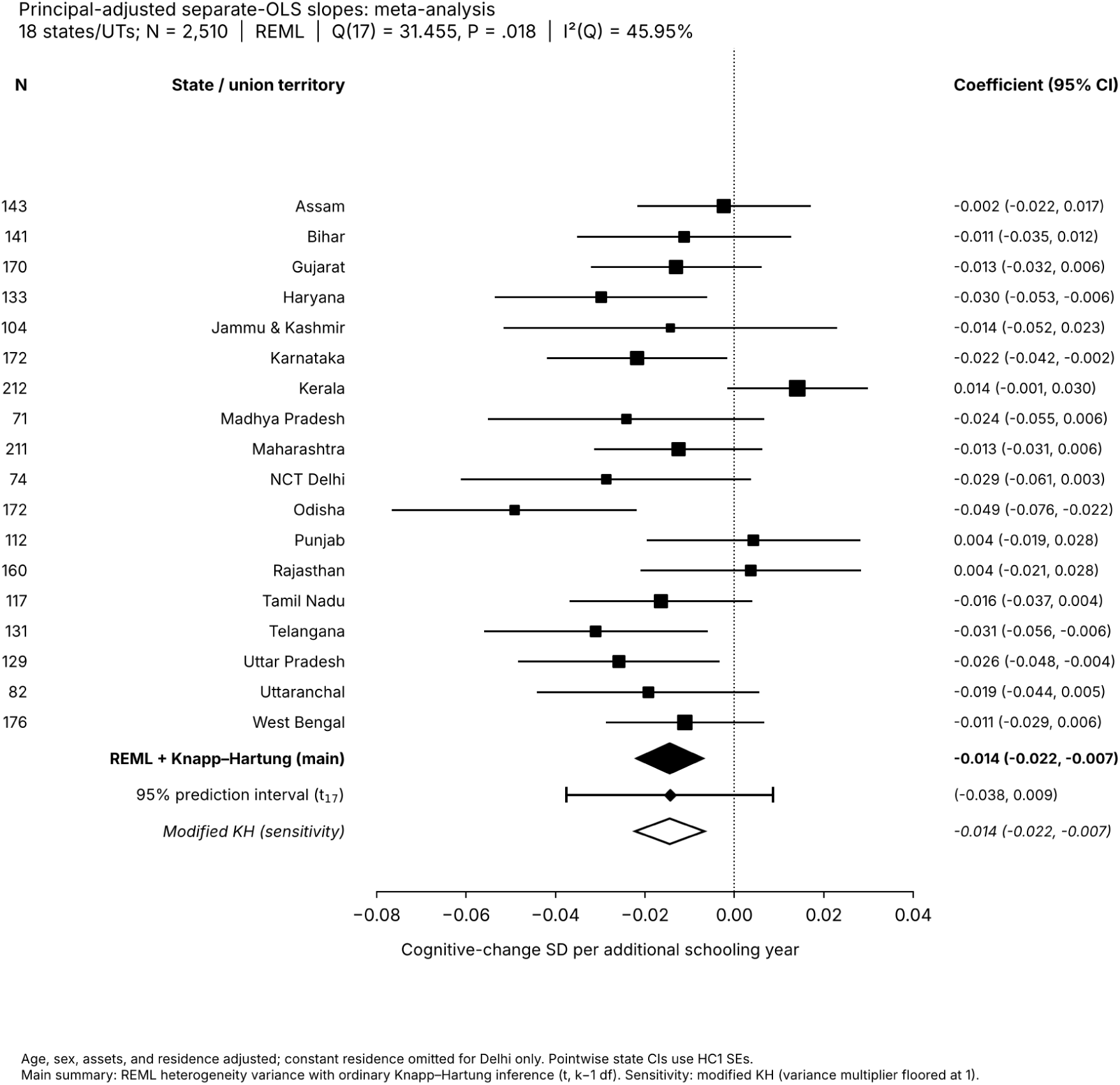
Forest plot of within-unit separate OLS slopes on the principal adjustment set, with the random-effects summary. *(REML estimator; pooled 95% CI and 95% prediction interval by the Knapp-Hartung method, t distribution with 17 degrees of freedom) and a modified Knapp-Hartung sensitivity interval.* *This separate-regression approach lets nuisance-covariate coefficients vary across units, unlike the pooled interaction model that shares them; the two are not interchangeable*.

**Supplementary Figure S3.**
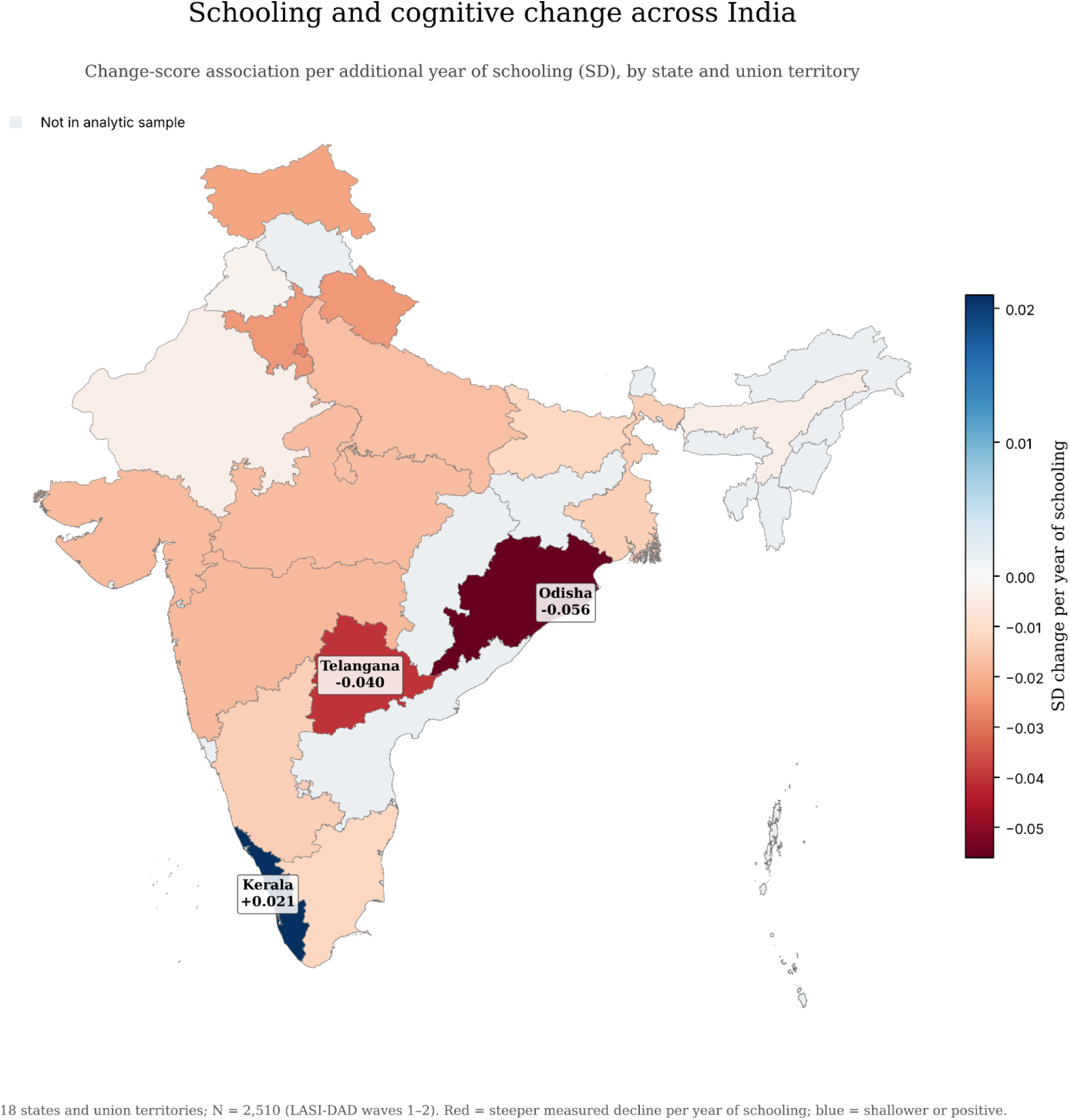
State- and union-territory-specific association between schooling and cognitive change across India. *Each shaded unit shows the change-score association per additional year of schooling, in SD units, from the principal model (N = 2,510; LASI-DAD waves 1–2). Red indicates a steeper measured decline per year of schooling; blue indicates a shallower or positive association. Units not in the analytic sample are shown in gray.* Boundaries are shown for illustration only and do not imply any position on disputed territories.

**Supplementary Table S1.** State- and union-territory-specific schooling associations with follow-up cognition (ANCOVA, conditional on baseline cognition)

| State / union territory | N | Coefficient | 95% CI | P value |
| --- | --- | --- | --- | --- |
| Assam | 143 | 0.048 | 0.032, 0.064 | <.001 |
| Bihar | 141 | 0.023 | 0.005, 0.040 | .012 |
| NCT Delhi | 74 | 0.000 | −0.022, 0.021 | .998 |
| Gujarat | 170 | 0.024 | 0.008, 0.039 | .003 |
| Haryana | 133 | 0.013 | −0.004, 0.030 | .130 |
| Jammu & Kashmir | 104 | 0.025 | 0.002, 0.048 | .030 |
| Karnataka | 172 | 0.029 | 0.014, 0.043 | <.001 |
| Kerala | 212 | 0.064 | 0.048, 0.079 | <.001 |
| Madhya Pradesh | 71 | 0.014 | −0.014, 0.042 | .330 |
| Maharashtra | 211 | 0.031 | 0.017, 0.045 | <.001 |
| Odisha | 172 | −0.009 | −0.029, 0.012 | .412 |
| Punjab | 112 | 0.022 | 0.006, 0.039 | .008 |
| Rajasthan | 160 | 0.041 | 0.018, 0.063 | <.001 |
| Tamil Nadu | 117 | 0.037 | 0.020, 0.054 | <.001 |
| Telangana | 131 | 0.000 | −0.018, 0.018 | .980 |
| Uttar Pradesh | 129 | 0.020 | 0.001, 0.039 | .041 |
| Uttaranchal | 82 | 0.016 | −0.004, 0.036 | .106 |
| West Bengal | 176 | 0.026 | 0.012, 0.039 | <.001 |

*Abbreviations: ANCOVA, analysis of covariance; CI, confidence interval; NCT, National Capital Territory. ANCOVA additionally conditions on baseline cognition; coefficients are the association of schooling with follow-up cognition given baseline, a different estimand from the change-score model, and are not an annual decline estimate. Estimates are adjusted for age, sex, household net assets, and rural or urban residence, with geography main effects.*
*Model N = 2,510. The robust schooling-by-geography omnibus test is $F(17, 2,469) = 4.11$ , $P = 3 \times 10^{-8}$ . The NCT Delhi estimate is −0.0000341 before rounding. Twelve intervals exclude zero and are positive; none is entirely negative.*

**Supplementary Table S2.**
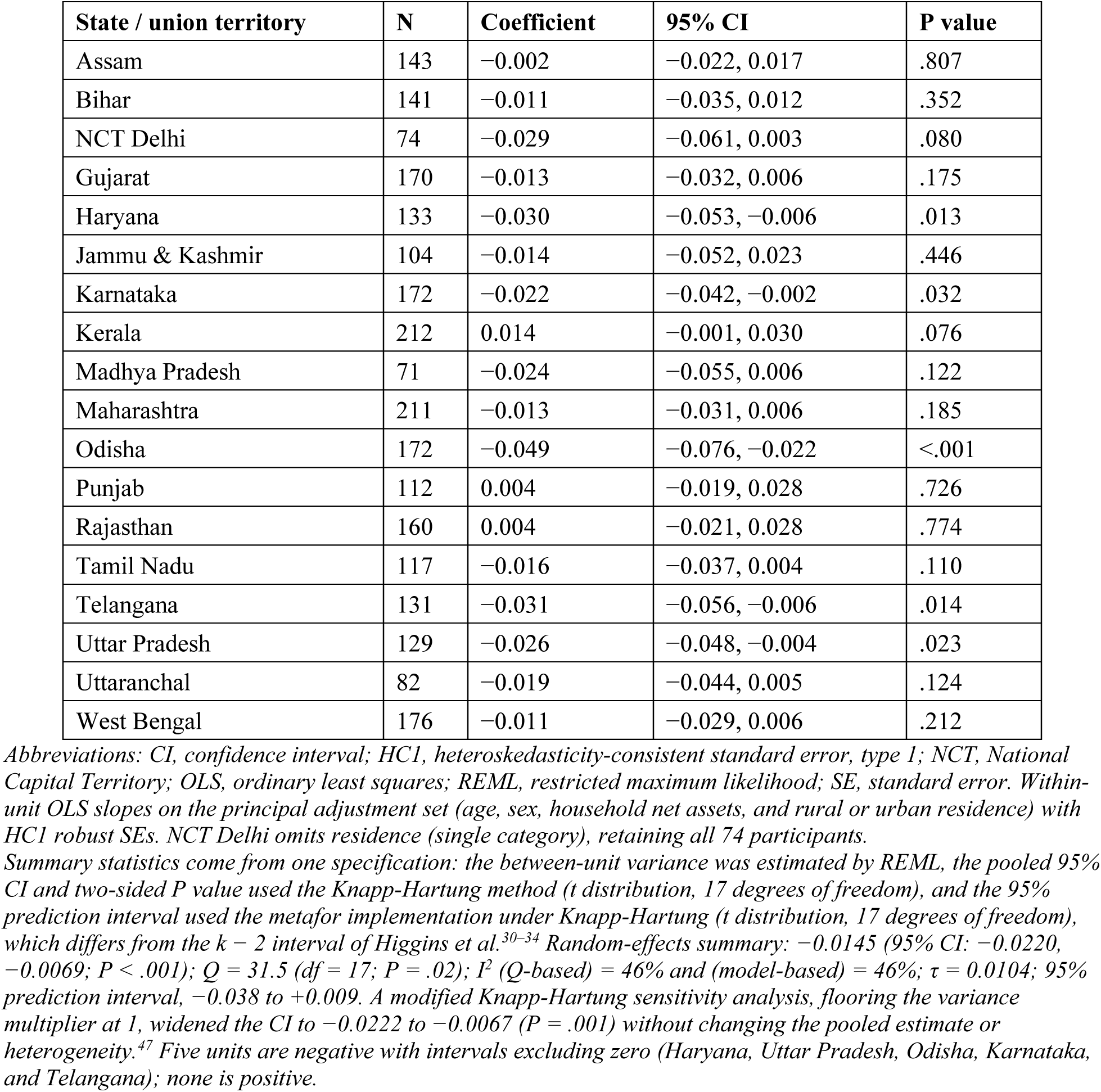
Within-unit separate ordinary least squares slopes (principal adjustment set) with random-effects summary.

**Supplementary Table S3.**
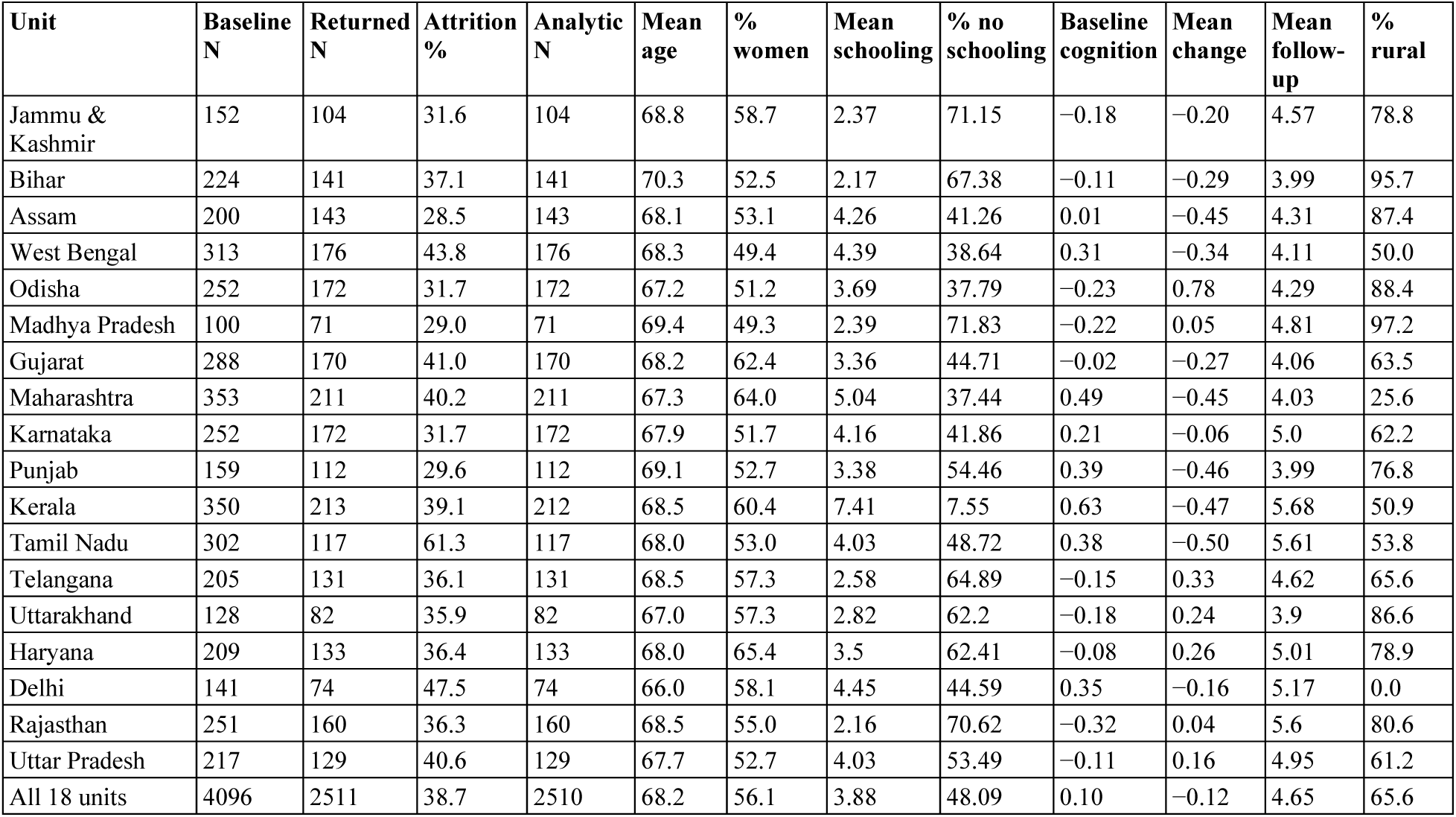
Unit-specific sample size, attrition, and baseline characteristics, LASI-DAD waves 1–2 (2017–2024).

| Unit | Baseline N | Returned N | Attrition % | Analytic N | Mean age | % women | Mean schooling | % no schooling | Baseline cognition | Mean change | Mean follow-up | % rural |
| --- | --- | --- | --- | --- | --- | --- | --- | --- | --- | --- | --- | --- |
| Jammu & Kashmir | 152 | 104 | 31.6 | 104 | 68.8 | 58.7 | 2.37 | 71.15 | −0.18 | −0.20 | 4.57 | 78.8 |
| Bihar | 224 | 141 | 37.1 | 141 | 70.3 | 52.5 | 2.17 | 67.38 | −0.11 | −0.29 | 3.99 | 95.7 |
| Assam | 200 | 143 | 28.5 | 143 | 68.1 | 53.1 | 4.26 | 41.26 | 0.01 | −0.45 | 4.31 | 87.4 |
| West Bengal | 313 | 176 | 43.8 | 176 | 68.3 | 49.4 | 4.39 | 38.64 | 0.31 | −0.34 | 4.11 | 50.0 |
| Odisha | 252 | 172 | 31.7 | 172 | 67.2 | 51.2 | 3.69 | 37.79 | −0.23 | 0.78 | 4.29 | 88.4 |
| Madhya Pradesh | 100 | 71 | 29.0 | 71 | 69.4 | 49.3 | 2.39 | 71.83 | −0.22 | 0.05 | 4.81 | 97.2 |
| Gujarat | 288 | 170 | 41.0 | 170 | 68.2 | 62.4 | 3.36 | 44.71 | −0.02 | −0.27 | 4.06 | 63.5 |
| Maharashtra | 353 | 211 | 40.2 | 211 | 67.3 | 64.0 | 5.04 | 37.44 | 0.49 | −0.45 | 4.03 | 25.6 |
| Karnataka | 252 | 172 | 31.7 | 172 | 67.9 | 51.7 | 4.16 | 41.86 | 0.21 | −0.06 | 5.0 | 62.2 |
| Punjab | 159 | 112 | 29.6 | 112 | 69.1 | 52.7 | 3.38 | 54.46 | 0.39 | −0.46 | 3.99 | 76.8 |
| Kerala | 350 | 213 | 39.1 | 212 | 68.5 | 60.4 | 7.41 | 7.55 | 0.63 | −0.47 | 5.68 | 50.9 |
| Tamil Nadu | 302 | 117 | 61.3 | 117 | 68.0 | 53.0 | 4.03 | 48.72 | 0.38 | −0.50 | 5.61 | 53.8 |
| Telangana | 205 | 131 | 36.1 | 131 | 68.5 | 57.3 | 2.58 | 64.89 | −0.15 | 0.33 | 4.62 | 65.6 |
| Uttarakhand | 128 | 82 | 35.9 | 82 | 67.0 | 57.3 | 2.82 | 62.2 | −0.18 | 0.24 | 3.9 | 86.6 |
| Haryana | 209 | 133 | 36.4 | 133 | 68.0 | 65.4 | 3.5 | 62.41 | −0.08 | 0.26 | 5.01 | 78.9 |
| Delhi | 141 | 74 | 47.5 | 74 | 66.0 | 58.1 | 4.45 | 44.59 | 0.35 | −0.16 | 5.17 | 0.0 |
| Rajasthan | 251 | 160 | 36.3 | 160 | 68.5 | 55.0 | 2.16 | 70.62 | −0.32 | 0.04 | 5.6 | 80.6 |
| Uttar Pradesh | 217 | 129 | 40.6 | 129 | 67.7 | 52.7 | 4.03 | 53.49 | −0.11 | 0.16 | 4.95 | 61.2 |
| All 18 units | 4096 | 2511 | 38.7 | 2510 | 68.2 | 56.1 | 3.88 | 48.09 | 0.10 | −0.12 | 4.65 | 65.6 |

**Supplementary Table S4.** Sensitivity of the schooling-by-geography interaction tests to follow-up length, standard-error type, small units, and unit-specific covariate coefficients.

| Interaction test (schooling × geography) | F | df1 | df2 | P | N |
| --- | --- | --- | --- | --- | --- |
| Change score, principal (HC1) | 2.97 | 17 | 2470 | $4.1 \times 10^{-5}$ | 2,510 |
| ANCOVA, principal (HC1) | 4.11 | 17 | 2469 | $3.1 \times 10^{-6}$ | 2,510 |
| Change score, principal (HC3) | 2.87 | 17 | 2470 | $7.2 \times 10^{-5}$ | 2,510 |
| ANCOVA, principal (HC3) | 3.96 | 17 | 2469 | $8.0 \times 10^{-6}$ | 2,510 |
| Change score + follow-up interval | 2.92 | 17 | 2469 | $5.4 \times 10^{-5}$ | 2,510 |
| Change score + schooling × interval | 3.40 | 17 | 2468 | $2.9 \times 10^{-5}$ | 2,510 |
| Annualized change outcome | 3.18 | 17 | 2470 | $1.2 \times 10^{-5}$ | 2,510 |
| ANCOVA + follow-up interval | 4.07 | 17 | 2468 | $4.0 \times 10^{-8}$ | 2,510 |
| Change, excluding units with N < 100 | 3.42 | 14 | 2249 | $1.6 \times 10^{-4}$ | 2,283 |
| ANCOVA, excluding units with N < 100 | 4.46 | 14 | 2248 | $5.7 \times 10^{-7}$ | 2,283 |
| Change, unit-specific covariates | 2.67 | 17 | 2419 | $3.7 \times 10^{-4}$ | 2,510 |
| ANCOVA, unit-specific covariates | 3.21 | 17 | 2418 | $9.2 \times 10^{-5}$ | 2,510 |
| Schooling × caste interaction | 4.17 | 3 | 2490 | $5.9 \times 10^{-3}$ | 2,502 |

## REFERENCES

1. Livingston G, Huntley J, Liu KY, et al. Dementia prevention, intervention, and care: 2024 report of the Lancet standing Commission. The Lancet. 2024;404(10452):572–628. doi:10.1016/S0140-6736(24)01296-0

2. Lövdén M, Fratiglioni L, Glymour MM, Lindenberger U, Tucker-Drob EM. Education and Cognitive Functioning Across the Life Span. Psychol Sci Public Interest. 2020;21(1):6–41. doi:10.1177/1529100620920576

3. Zhang YS, O’Shea B, Yu X, et al. Educational Attainment and Later-Life Cognitive Function in High- and Middle-Income Countries: Evidence From the Harmonized Cognitive Assessment Protocol. Schafer M, ed. *The Journals of Gerontology*, Series B: Psychological Sciences and Social Sciences. 2024;79(5):gbae005. doi:10.1093/geronb/gbae005

4. Šeblová D, Berggren R, Lövdén M. Education and age-related decline in cognitive performance: systematic review and meta-analysis of longitudinal cohort studies. Ageing Research Reviews. 2020;58:101005. doi:10.1016/j.arr.2019.101005

5. Fjell AM, Rogeberg O, Sørensen Ø, et al. Reevaluating the role of education on cognitive decline and brain aging in longitudinal cohorts across 33 Western countries. Nat Med. 2025;31(9):2967–2976. doi:10.1038/s41591-025-03828-y

6. Zahodne LB, Glymour MM, Sparks C, et al. Education Does Not Slow Cognitive Decline with Aging: 12-Year Evidence from the Victoria Longitudinal Study. J Int Neuropsychol Soc. 2011;17(6):1039–1046. doi:10.1017/S1355617711001044

7. Stern Y. What is cognitive reserve? Theory and research application of the reserve concept. J Int Neuropsychol Soc. 2002;8(3):448–460. doi:10.1017/S1355617702813248

8. Stern Y. Cognitive reserve in ageing and Alzheimer’s disease. The Lancet Neurology. 2012;11(11):1006–1012. doi:10.1016/S1474-4422(12)70191-6

9. Lipnicki DM, Crawford JD, Dutta R, et al. Age-related cognitive decline and associations with sex, education and apolipoprotein E genotype across ethnocultural groups and geographic regions: a collaborative cohort study. Miller BL, ed. PLoS Med. 2017;14(3):e1002261. doi:10.1371/journal.pmed.1002261

10. Ruiz M, Hu Y, Martikainen P, Bobak M. Life Course Socioeconomic Position and Cognitive Aging Trajectories: A Cross-National Cohort Study in China and England. Albert SM, ed. Innovation in Aging. 2023;7(6):igad064. doi:10.1093/geroni/igad064

11. Orsholits D, Cullati S, Ghisletta P, et al. How Welfare Regimes Moderate the Associations Between Cognitive Aging, Education, and Occupation. Gutchess A, ed. The Journals of Gerontology: Series B. 2022;77(9):1615–1624. doi:10.1093/geronb/gbac013

12. Gross AL, Nichols E, Arce Rentería M, et al. Risk and protective factors for cognitive decline in older adults from a nationally representative sample in India: results from the LASI-DAD. Alzheimer’s & Dementia. 2026;22(7):e71602. doi:10.1002/alz.71602

13. Nichols E, Jones RN, Gross AL, et al. Exploring the association between higher education and steeper cognitive decline in a nationally representative longitudinal study in India. American Journal of Epidemiology. Published online January 15, 2026:kwag009. doi:10.1093/aje/kwag009

14. Lee J, Meijer E, Langa KM, et al. Prevalence of dementia in India: National and state estimates from a nationwide study. Alzheimer’s &amp; Dementia. 2023;19(7):2898–2912. doi:10.1002/alz.12928

15. Lee J, Smith JP. Regional disparities in adult height, educational attainment, and late-life cognition: Findings from the Longitudinal Aging Study in India (LASI). Journal of the Economics of Ageing. 2014;4:26–34. doi:10.1016/j.jeoa.2014.02.002

16. Walsemann KM, Ailshire JA. Early Educational Experiences and Trajectories of Cognitive Functioning Among US Adults in Midlife and Later. American Journal of Epidemiology. 2020;189(5):403–411. doi:10.1093/aje/kwz276

17. Walsemann KM, Jackson H, Abbruzzi E, Ailshire JA. State-Level Education Quality and Trajectories of Cognitive Function by Race and Educational Attainment. Milbank Quarterly. 2024;102(3):765–821. doi:10.1111/1468-0009.12709

18. Seblova D, Eng C, Avila-Rieger JF, et al. High school quality is associated with cognition 58 years later. Alz & Dem Diag Ass & Dis Mo. 2023;15(2):e12424. doi:10.1002/dad2.12424

19. Leist AK, Bar-Haim E, Chauvel L. Inequality of educational opportunity at time of schooling predicts cognitive functioning in later adulthood. SSM - Population Health. 2021;15:100837. doi:10.1016/j.ssmph.2021.100837

20. Lee J, Khobragade PY, Banerjee J, et al. Design and Methodology of the Longitudinal Aging Study in India-Diagnostic Assessment of Dementia (LASI-DAD). Journal of the American Geriatrics Society. 2020;68(S3):S5–S10. doi:10.1111/jgs.16737

21. Gross AL, Khobragade PY, Meijer E, Saxton JA. Measurement and Structure of Cognition in the Longitudinal Aging Study in India–Diagnostic Assessment of Dementia. J American Geriatrics Society. 2020;68(S3). doi:10.1111/jgs.16738

22. Lord FM. A paradox in the interpretation of group comparisons. Psychological Bulletin. 1967;68(5):304–305. doi:10.1037/h0025105

23. Van Breukelen GJP. ANCOVA Versus CHANGE From Baseline in Nonrandomized Studies: The Difference. Multivariate Behavioral Research. 2013;48(6):895–922. doi:10.1080/00273171.2013.831743

24. Glymour MM, Weuve J, Berkman LF, Kawachi I, Robins JM. When is baseline adjustment useful in analyses of change? An example with education and cognitive change. American Journal of Epidemiology. 2005;162(3):267–278. doi:10.1093/aje/kwi187

25. Tennant PWG, Arnold KF, Ellison GTH, Gilthorpe MS. Analyses of ‘change scores’ do not estimate causal effects in observational data. International Journal of Epidemiology. 2022;51(5):1604–1615. doi:10.1093/ije/dyab050

26. Khobragade PY, Petrosyan S, Dey S, Dey AB, Lee J. Design and methodology of the harmonized Diagnostic Assessment of Dementia for the Longitudinal Aging Study in India: Wave 2. Journal of the American Geriatrics Society. 2025;73(3):685–696. doi:10.1111/jgs.19252

27. Gross AL, Li C, Briceño EM, et al. Harmonisation of later-life cognitive function across national contexts: results from the Harmonized Cognitive Assessment Protocols. The Lancet Healthy Longevity. 2023;4(10):e573–e583. doi:10.1016/S2666-7568(23)00170-8

28. Stram DO, Lee JW. Variance Components Testing in the Longitudinal Mixed Effects Model. Biometrics. 1994;50(4):1171. doi:10.2307/2533455

29. Higgins JPT, Thompson SG. Quantifying heterogeneity in a meta-analysis. Statistics in Medicine. 2002;21(11):1539–1558. doi:10.1002/sim.1186

30. Knapp G, Hartung J. Improved tests for a random effects meta-regression with a single covariate. Statistics in Medicine. 2003;22(17):2693–2710. doi:10.1002/sim.1482

31. IntHout J, Ioannidis JP, Borm GF. The Hartung-Knapp-Sidik-Jonkman method for random effects meta-analysis is straightforward and considerably outperforms the standard DerSimonian-Laird method. BMC Med Res Methodol. 2014;14(1):25. doi:10.1186/1471-2288-14-25

32. Higgins JPT, Thompson SG, Spiegelhalter DJ. A re-evaluation of random-effects meta-analysis. Journal of the Royal Statistical Society: Series A (Statistics in Society). 2009;172(1):137–159. doi:10.1111/j.1467-985X.2008.00552.x

33. Riley RD, Higgins JPT, Deeks JJ. Interpretation of random effects meta-analyses. BMJ. 2011;342(feb10 2):d549-d549. doi:10.1136/bmj.d549

34. IntHout J, Ioannidis JPA, Rovers MM, Goeman JJ. Plea for routinely presenting prediction intervals in meta-analysis. BMJ Open. 2016;6(7):e010247. doi:10.1136/bmjopen-2015-010247

35. Weuve J, Tchetgen Tchetgen EJ, Glymour MM, et al. Accounting for bias due to selective attrition: the example of smoking and cognitive decline. Epidemiology. 2012;23(1):119–128. doi:10.1097/EDE.0b013e318230e861

36. Solon G, Haider SJ, Wooldridge JM. What are we weighting for? Journal of Human Resources. 2015;50(2):301–316. doi:10.3368/jhr.50.2.301

37. Bates D, Mächler M, Bolker B, Walker S. Fitting Linear Mixed-Effects Models Using **lme4**. J Stat Soft. 2015;67(1). doi:10.18637/jss.v067.i01

38. Viechtbauer W. Conducting Meta-Analyses in *R* with the **metafor** Package. J Stat Soft. 2010;36(3). doi:10.18637/jss.v036.i03

39. Nichols E, Dey S, Khobragade P, et al. Socioeconomic and clinical predictors of all-cause mortality among older adults in India from 2017 to 2024: the importance of cognitive health and dementia. The Journals of Gerontology: Series A. 2026;81(3):glag026. doi:10.1093/gerona/glag026

40. Srikantha S, Manne-Goehler J, Kobayashi LC, Flood D, Koton S, Gross AL. Type II diabetes and cognitive function among older adults in India and China: results from Harmonized Cognitive Assessment Protocol studies. Frontiers in Public Health. 2024;12:1474593. doi:10.3389/fpubh.2024.1474593

41. Clouston SAP, Smith DM, Mukherjee S, et al. Education and Cognitive Decline: An Integrative Analysis of Global Longitudinal Studies of Cognitive Aging. Brown JS, ed. The Journals of Gerontology: Series B. 2020;75(7):e151–e160. doi:10.1093/geronb/gbz053

42. Mungas D, Gavett B, Fletcher E, Farias ST, DeCarli C, Reed B. Education amplifies brain atrophy effect on cognitive decline: implications for cognitive reserve. Neurobiology of Aging. 2018;68:142–150. doi:10.1016/j.neurobiolaging.2018.04.002

43. Crowe M, Clay OJ, Martin RC, et al. Indicators of Childhood Quality of Education in Relation to Cognitive Function in Older Adulthood. The Journals of Gerontology Series A: Biological Sciences and Medical Sciences. 2013;68(2):198–204. doi:10.1093/gerona/gls122

44. Goel A, Gross A. Differential item functioning in the cognitive screener used in the Longitudinal Aging Study in India. International Psychogeriatrics. 2019;31(9):1331–1341. doi:10.1017/S1041610218001746

45. Nichols E, Ng DK, Hayat S, et al. Differences in the measurement of cognition for the assessment of dementia across geographic contexts: Recommendations for cross-national research. Alzheimer’s &amp; Dementia. 2023;19(3):1009–1019. doi:10.1002/alz.12740

46. Liu Y, Lynch KM, Arce Rentería M, et al. Superficial white matter and gray matter jointly support cognition among older adults in India. Alzheimer’s & Dementia. 2026;22(7):e71697. doi:10.1002/alz.71697

47. Röver C, Knapp G, Friede T. Hartung-Knapp-Sidik-Jonkman approach and its modification for random-effects meta-analysis with few studies. BMC Med Res Methodol. 2015;15(1):99. doi:10.1186/s12874-015-0091-1

